# Clinical Characteristics of Patients with Severe Pneumonia Caused by the 2019 Novel Coronavirus in Wuhan, China

**DOI:** 10.1101/2020.03.02.20029306

**Authors:** Yafei Wang, Ying Zhou, Zhen Yang, Dongping Xia, Shuang Geng

## Abstract

**Background:** A new virus broke out in Wuhan, Hubei, China, and was later named 2019 novel coronavirus (2019-nCoV). The clinical characteristics of severe pneumonia caused by 2019-nCoV are still not clear.

**Objectives:** The aim of this study was to explore the clinical characteristics and risk factors of the severe pneumonia caused by the 2019-nCoV in Wuhan, China.

**Method:** The study included patients hospitalized at the central hospital of Wuhan who had been diagnosed with a pneumonia caused by the novel coronavirus. Clinical features, chronic co-morbidities, demographic data, laboratory examinations, and chest computed tomography (CT) scans were reviewed through electronic medical records. SPSS was used for data analysis to explore the clinical characteristics and risk factors of the patients with the severe pneumonia.

**Results:** A total of 110 patients diagnosed with 2019 novel coronavirus pneumonia were included in the study, including 38 with severe pneumonia and 72 with non-severe pneumonia. Statistical analysis showed that advanced age, an increase of D-dimer, and a decrease of lymphocytes were characteristics of the patients with severe pneumonia. Moreover, in the early stage of the disease, chest CT scans of patients with the severe pneumonia showed the illness can progress rapidly.

**Conclusions:** Advanced age, lymphocyte decline, and D-dimer elevation are important characteristics of patients with severe pneumonia. Clinicians should focus on these characteristics to identify high-risk patients at an early stage.

## Introduction

In December 2019, a new type of unexplained pneumonia was reported in Wuhan, Hubei, China, which appeared to be related to the Huanan Seafood Wholesale Market[1-3]. The disease spread rapidly from Wuhan to the surrounding provinces and cities, which got the attention of the government and the administrative departments of health at all levels. The Chinese Center for Disease Control and Prevention (CDC) promptly organized the relevant disease control agencies, medical units, and research institutes to carry out investigations and treatment. A new type of coronavirus was detected by researchers in a patient’s bronchoalveolar lavage fluid sample on January 3, 2020[4]. The World Health Organization (WHO) named it the 2019-novel coronavirus (2019-nCoV) and announced that the new coronavirus epidemic had been listed as a public health emergency of international concern on January 30, 2020. As of 18:00 on February 11, 2020, there were 42,744 confirmed cases, 21,675 suspected cases, 4,161 cured cases, and 1,017 deaths in China.

The 2019-nCoV, which belongs to the genus betacoronavirus, is a single-stranded positive-strand RNA virus that appears to be distinct from, but is related to, other coronaviruses, such as severe acute respiratory syndrome-related coronavirus (SARSr-CoV) and Middle East respiratory syndrome coronavirus (MERSr-CoV)[4-8]. Current studies have shown that 2019-nCoV has about an 89% homology with bat SARS-like-CoVZXC21 and 82% homology with human SARS-CoV[9]. The disease is highly contagious, and may rapidly develop into severe pneumonia, acute respiratory distress syndrome (ARDS), multiple organ dysfunction syndrome (MODS), and death, so the top priority for clinicians is to identify and treat the severest patients in the early stage. The aim of this study was to assess the potential high-risk factors of 2019-nCoV severe pneumonia and provide evidence for the screening of severely afflicted patients.

## Methods

### Patients

All of the patients in this study were hospitalized in a respiratory department, the Respiratory Intensive Care Unit (RICU), from January 1, 2020 to February 10, 2020. The patients were all admitted to the hospital because they were infected with 2019-nCoV and suffered from various kinds of symptoms, including fever, dyspnea, cough, and fatigue. Every patient had completed the relevant laboratory examination, including various common pathogen detection and chest computed tomographic (CT) scans. All of the patients were local residents of Wuhan. Moreover, most of the patients had a history of exposure to the Huanan Seafood Wholesale Market or had made contact with people who had been confirmed (or suspected) to have contracted the illness. The 2019-nCoV nucleic acid detection of some patients was positive, while for others, it was negative. These results could have been caused by the immaturity of the methods used for 2019-nCoV nucleic acid detection, which led to false negative results. High-resolution CT scans with a scan layer thickness of 5 mm and a reconstruction of a 1-1.5 mm thin layer are recommended for the radiological examination of 2019n-CoV pneumonia. Based on the patients’ exposure history, clinical symptoms, laboratory examinations, and chest CT scans, all of the patients were clinically diagnosed with 2019-nCoV pneumonia according to WHO’s interim guidance[10]. For patients who were suspected to have the illness, two senior respiratory doctors made the diagnosis together.

The patients were divided into two groups: patients with severe pneumonia and those with non-severe pneumonia. The former referred to patients with the following severe manifestations: fever or suspected respiratory infection, plus one of a respiratory rate >30 breaths/min, severe respiratory distress, or SpO2 <90% on room air. Patients with ARDS, sepsis, or septic shock were also included. The patients without the above severe signs were defined as having non-severe pneumonia. Patients who had the illness combined with other bacterial, fungal, or other viral infections and those with missing data were excluded. The study was approved by the ethics committee of Wuhan Central Hospital (Yuan lun han (2020) no.4).

### Data collection

The patient data were extracted from the Central Hospital of Wuhan, which is a tertiary teaching hospital and is responsible for the treatment for patients with 2019-nCoV pneumonia, as assigned by the Chinese government. Clinical features, chronic co-morbidities, demographic data, laboratory examinations, and chest CT scans were reviewed using electronic medical records. Laboratory examinations included routine blood tests, as well as tests of the liver function, kidney function, electrolytes, B-type natriuretic peptide, D-dimer, C-reactive protein, and procalcitonin. We obtained the lymphocyte absolute values of patients with severe pneumonia on the first day and the third day after admission. The D-dimer data of patients with severe pneumonia on the first day, the third day, and the seventh day after admission were also collected. Chest CT scans were reviewed on the first day and the third day for patients with severe pneumonia. In addition to collecting the albumin data on the first day after admission, the data on the seventh day were also obtained for patients with severe pneumonia. For patients with non-severe pneumonia, we only collected the data on the first day after admission because the relevant items were not frequently reviewed. For patients admitted to the RICU, the Acute Physiology and Chronic Health Evaluation II scores (APACHE-II) and Sequential Organ Failure Assessment (SOFA) were determined on the first day. The data were acquired by physicians. All of the data were checked by another researcher to ascertain its accuracy. To reflect the progression of the disease in critically ill patients, we calculated the difference in the lymphocyte values between day 3 and day 1, the difference in the serum albumin values between day 7 and day 1, and the difference in the D-dimer values between days 3, 7, and 1.

### Statistical analysis

The continuous variables were used as the mean and compared using the t-tests if they were normally distributed, or they were described using the median. The Mann-Whitney U test was used for comparisons. Categorical variables were expressed as count (%) and compared by χ^2^ test or Fisher’s exact test. Logistic regression analysis was used to assess the risk factors of severe pneumonia. The difference of a certain indicator in the same patient at different periods was shown by a bar chart. A two-sided α of less than 0.05 was considered statistically significant. We used SPSS software (version 23.0) for statistical analysis.

## Results

### Basic characteristics

A total of 110 hospitalized patients participated in this study, which included 38 (34.5%) patients with severe pneumonia and 72 (65.5%) patients with non-severe pneumonia. Table 1 shows that compared with patients with non-severe pneumonia, males accounted for a greater proportion of those with severe pneumonia and the difference was significant (24 [63.16%] vs. 24 [33.33%]). The patients with severe pneumonia tended to be older and had complications with COPD (4 [10.53%] vs. 2 [2.78%]) and hypertension (15 [39.47%] vs. 8 [11.11%]). Patients over the age of 60 years accounted for a greater proportion of the severe pneumonia cases (27 [71.05%] vs. 9 [12.5%]). There was no significant difference in the smoking history and drinking history between the two groups. The incidence of diabetes and cerebrovascular disease was similar in both groups. According to the patients’ medical history, the common symptoms at the onset of the illness were fever, fatigue, dry cough, and dyspnea. Although the initial symptoms of the patients with severe pneumonia were more commonly fever and dyspnea, and the difference was not statistically significant. The temperature ranges were divided into low fever (temperature ≤38°C), moderate fever (temperature ≥38.1°C, ≤39°C), and high fever (temperature ≥39.1°C). However, there was no statistical difference between the two groups.

**Table 1.** Characteristics of patients with severe and non-severe pneumonia

|  | Normal Range | Severe Pneumonia (n=38) | Non-severe Pneumonia (n = 72) | P Value |
| --- | --- | --- | --- | --- |
| Male (%) | NA | 24(63.16) | 24(33.33) | 0.004 |
| Age range (years) | NA |  |  | <0.001 |
| ≤40 |  | 3 (7.89%) | 50(69.44%) |  |
| 40<, ≤60 |  | 8(21.05%) | 13(18.06%) |  |
| ≥60 |  | 27(71.05) | 9(12.5%) |  |
| Fever (%) | NA | 33 (86.84%) | 58 (79.17%) | 0.438 |
| Highest temperature (%) | NA |  |  | 0.103 |
| Low fever |  | 5(13.16%) | 22(30.56%) |  |
| Moderate fever |  | 25(65.79%) | 31(43.06%) |  |
| High fever |  | 3(7.89%) | 5(6.94%) |  |
| Dyspnea (%) | NA | 15(39.47%) | 21(29.17%) | 0.292 |
| Dry cough (%) | NA | 21(55.26%) | 47(65.28%) | 0.312 |
| Fatigue (%) | NA | 10(26.32%) | 31(43.06%) | 0.100 |
| COPD (%) | NA | 4(10.53%) | 2(2.78%) | 0.013 |
| Diabetes (%) | NA | 8(21.05%) | 7(9.72%) | 0.143 |
| Hypertension (%) | NA | 15(39.47%) | 8(11.11%) | 0.001 |
| Cerebrovascular disease (%) | NA | 3(7.89%) | 4(5.56%) | 0.691 |
| Smoking (%) | NA | 9(23.68%) | 17(23.61%) | 0.96 |
| Drinking (%) | NA | 4(10.53%) | 19(26.39%) | 0.083 |
| White blood cell count (10<sup>9</sup>/L) | 3.5-9.5 | 5.20(3.90,6.46) | 5.21(4.11,6.80) | 0.772 |
| Neutrophil count (10<sup>9</sup>/L) | 1.8-6.3 | 4.26(2.84,4.84) | 3.38(2.33,5.24) | 0.258 |
| Lymphocyte count (10<sup>9</sup>/L) | 1.1-3.2 | 0.60(0.31) | 1.21(0.53) | <0.001 |
| Platelet count (10<sup>9</sup>/L) | 125-350 | 144.50(110.75,167.75) | 179.5(151.75,226.50) | <0.001 |
| Hemoglobin (g/L) | 130-175 | 129.87(19.39) | 132.43(16.07) | 0.461 |
| Serum creatinine (umol/L) | 57-111 | 74.75(59.10,93.10) | 55.55(43.73,72.00) | <0.001 |
| Blood Urea Nitrogen (mmol/L) | 2.78-8.07 | 4.99(3.97,6.53) | 3.93(3.19,4.74) | <0.001 |
| Alanine aminotransferase (U/L) | 9-50 | 21.90(17.50,36.83) | 19.75(14.45,32.10) | 0.304 |
| Aspartate aminotransferase (U/L) | 10-60 | 36.80(27.85,45.00) | 20.00(15.85,26.40) | <0.001 |
| Serum albumin (g/L) | 40-55 | 35.30(4.91) | 40.97(4.42) | <0.001 |
| C-reactive protein (mg/dL) | <0.5 | 4.98(2.72,9.74) | 0.52(0.13,2.65) | <0.001 |
| Serum procalcitonin (ng/ml) | <0.5 | 0.12(0.06,0.33) | 0.05(0.04,0.10) | <0.001 |
| D - dimer (ug/ml.FEU) | <1 | 1.11(0.47,3.83) | 0.37(0.21,0.78) | <0.001 |
| B-type natriuretic peptide (pg/ml) | <125 | 134.60(87.25,394.70) | 43.50(19.00,80.50) | <0.001 |

Age was divided into three ranges: ≤40 years; 40 years<, ≤60 years; ≥60 years
The temperature ranges were divided into low (≤38°C), moderate (≥38.1°C, ≤39°C), and high fevers (≥39.1°C).

### Laboratory parameters

There was a number of differences in the laboratory findings between the patients with severe pneumonia and those without it (Table 1), including a lower lymphocyte count, platelet count, and serum albumin. The level of serum creatinine, blood urea nitrogen, aspartate aminotransferase, c-reactive protein, serum procalcitonin, D-dimer, and B-type natriuretic peptide were higher in the patients with severe pneumonia. The white blood cell count, neutrophil count, hemoglobin, and alanine aminotransferase did not differ between the two groups. Statistically, some parameters were different between the two groups, but still within the normal range, including the platelet count, serum creatinine, blood urea nitrogen, aspartate aminotransferase, and serum procalcitonin.

### Characteristics of patients with severe pneumonia

Binomial logistics regression analysis was used to assess the risk factors of severe pneumonia. The variables with statistical differences between the two groups were incorporated into in the logistics regression equation. To better understand the correlation between the lymphocytes and D-dimer measurements with severe pneumonia, we divided the values of the lymphocytes and d-dimer by their standard deviations. The results revealed that after adjusting for other confounding factors, the age and D-dimer values were independent risk factors of severe pneumonia (Table 2). Patients over the age of 60 years and in the range of 40 to 60 old had a significantly higher risk of developing severe pneumonia than those under the age of 40. For every 1 standard deviation increase in the D-dimer value, the patients’ risk of developing severe pneumonia increased by about 17 times. In addition, lymphocytes were found to be independent protective factors for severe pneumonia. The incidence of severe pneumonia in patients with a 0.55 increase in lymphocytes decreased by about 67.8%.

**Table 2.**
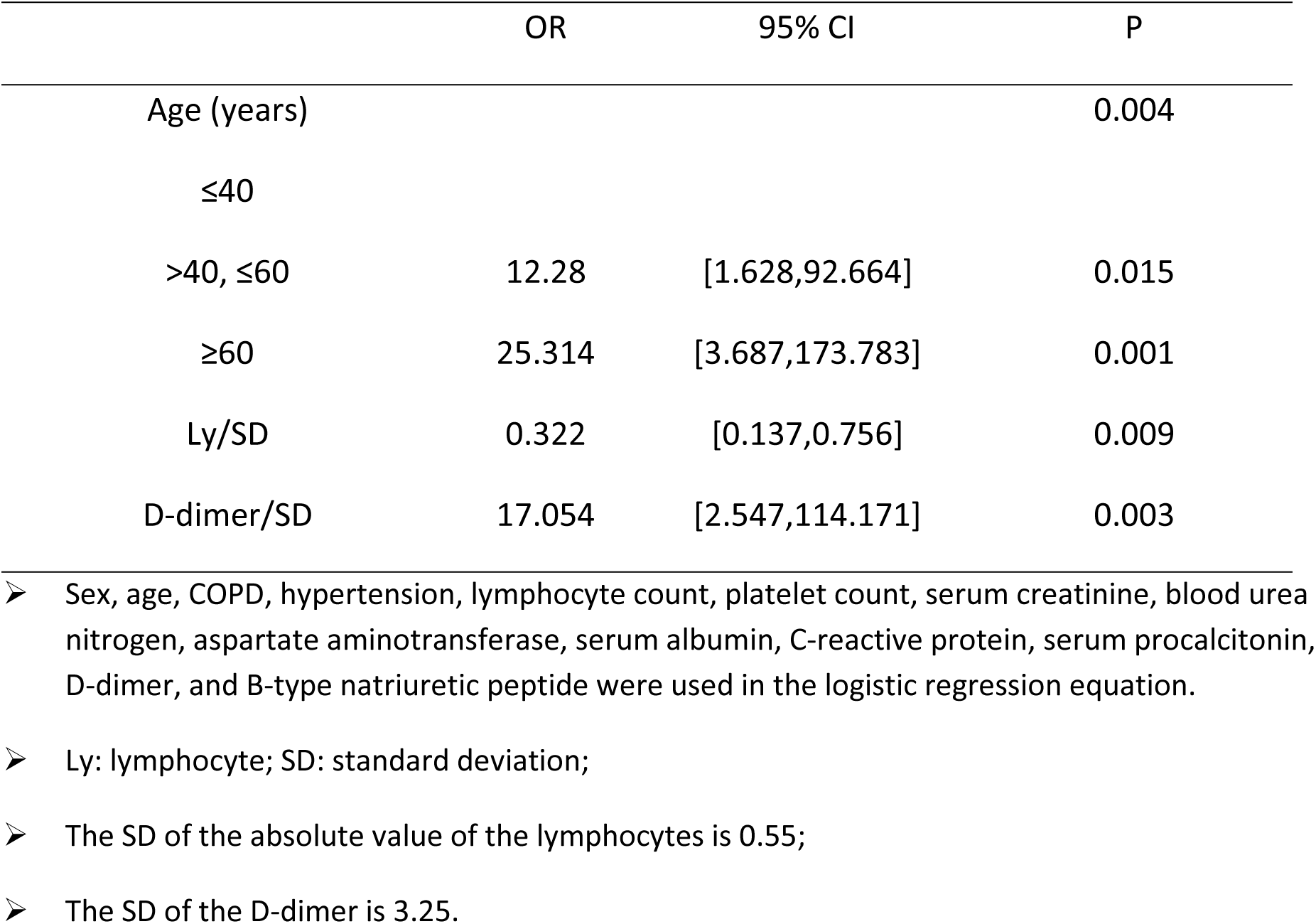
The risk factors of 2019 Novel Coronavirus Severe Pneumonia

Both the APACHE-II and SOFA scores were assessed in patients with severe pneumonia, with results of 14.5 (13, 17) and 6 (5, 7), respectively (Table 3). Severe pneumonia usually progresses rapidly, and many clinical indicators can change in a short time, especially the lymphocyte, D-dimer, serum albumin, and chest CT manifestations. To better identify severe pneumonia early, we calculated the difference in the lymphocyte counts, serum albumin values, and D-dimer values at different points in time (Table 3, Figs. 1-4). The results showed that in the early stage of the disease, the lymphocyte and serum albumin decreased and the D-dimer increased with the progress of the disease. We have included 2 chest CT scans of one patient with severe pneumonia at different time periods, which suggested the rapid progress of the disease (Fig. 5).

**Table 3.**
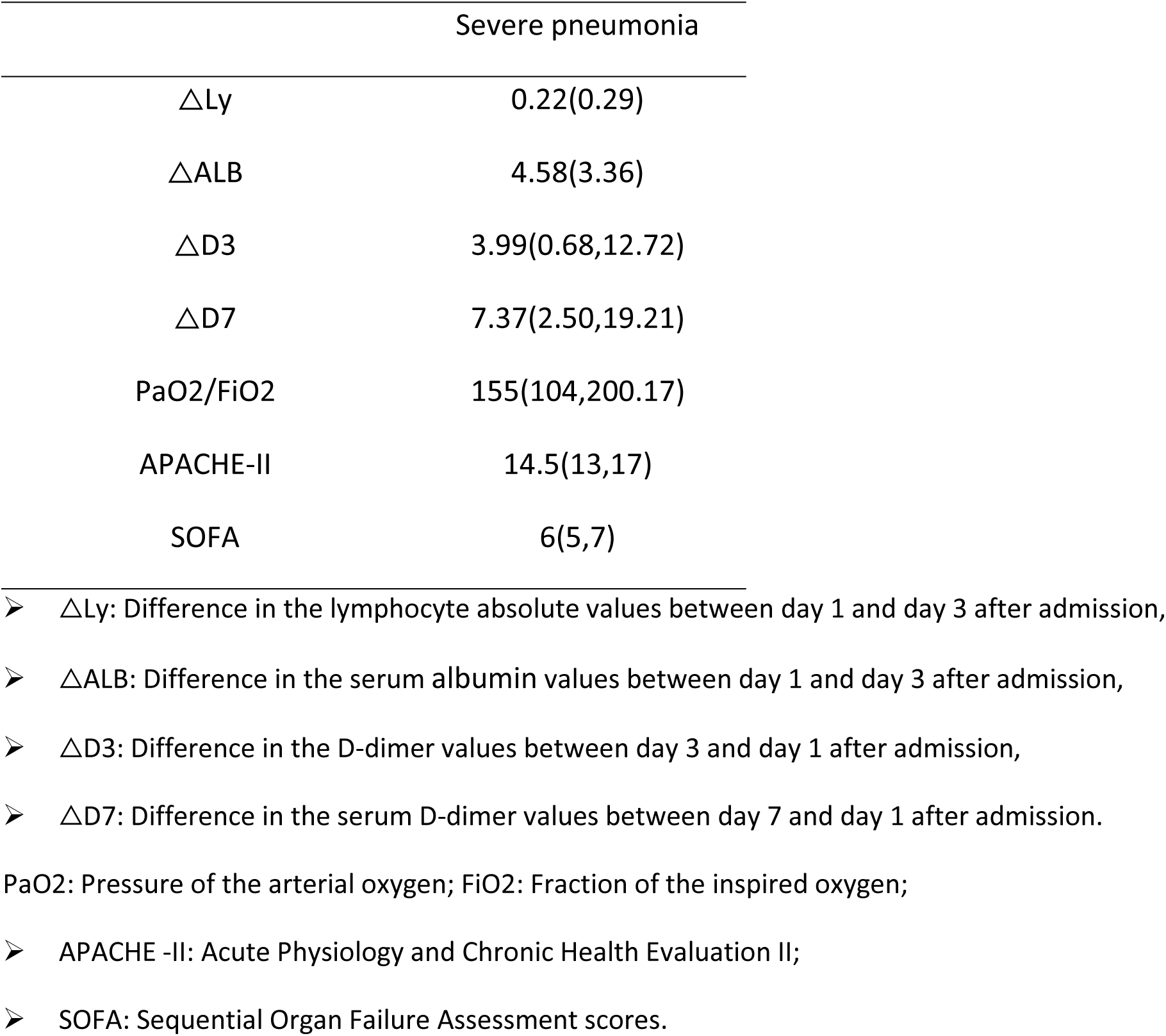
Characteristics of patients with severe pneumonia

|  | Severe pneumonia |
| --- | --- |
| ΔLy | 0.22(0.29) |
| ΔALB | 4.58(3.36) |
| ΔD3 | 3.99(0.68,12.72) |
| ΔD7 | 7.37(2.50,19.21) |
| PaO<sub>2</sub>/FiO<sub>2</sub> | 155(104,200.17) |
| APACHE-II | 14.5(13,17) |
| SOFA | 6(5,7) |

- $\Delta$ Ly: Difference in the lymphocyte absolute values between day 1 and day 3 after admission, - $\Delta$ ALB: Difference in the serum albumin values between day 1 and day 3 after admission, - $\Delta$ D3: Difference in the D-dimer values between day 3 and day 1 after admission, - $\Delta$ D7: Difference in the serum D-dimer values between day 7 and day 1 after admission.
PaO<sub>2</sub>: Pressure of the arterial oxygen; FiO<sub>2</sub>: Fraction of the inspired oxygen;
- APACHE -II: Acute Physiology and Chronic Health Evaluation II; - SOFA: Sequential Organ Failure Assessment scores.

**Figure 1.**
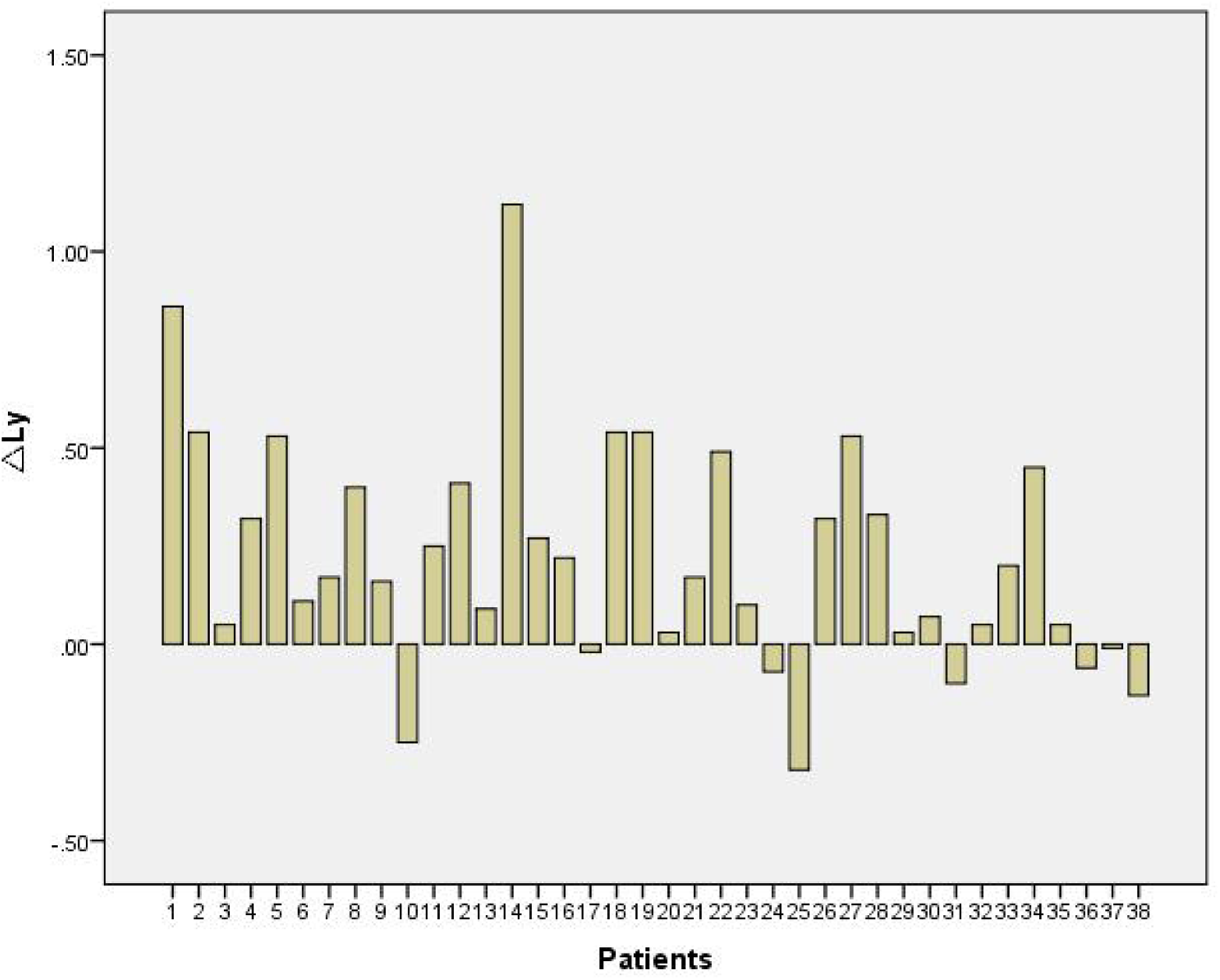
△Ly: Difference in the lymphocyte absolute values between day 1 and day 3 after admission

**Figure 2.**
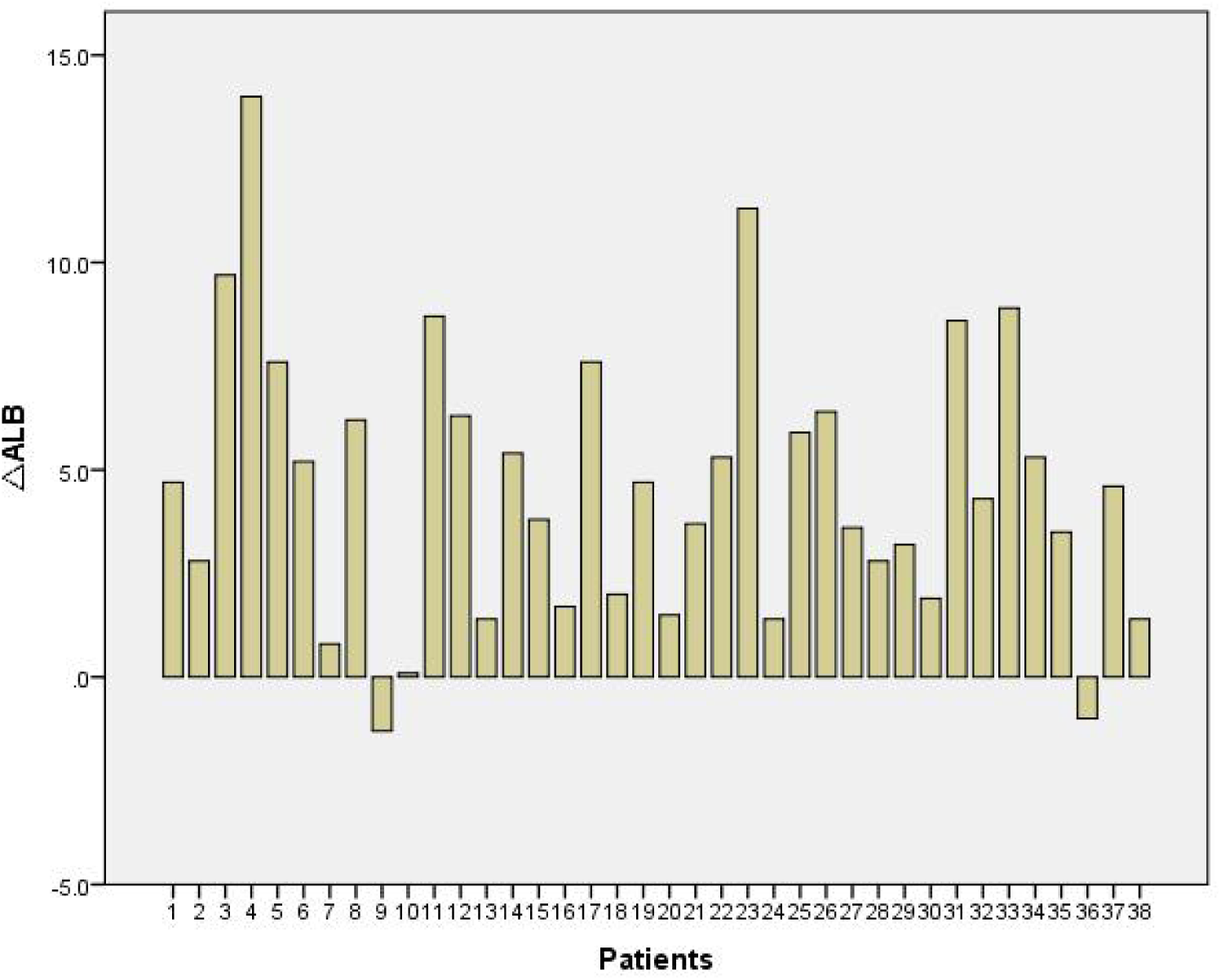
△ALB:Difference in the serum albumin values between day 1 and day 3 after admission

**Figure 3.**
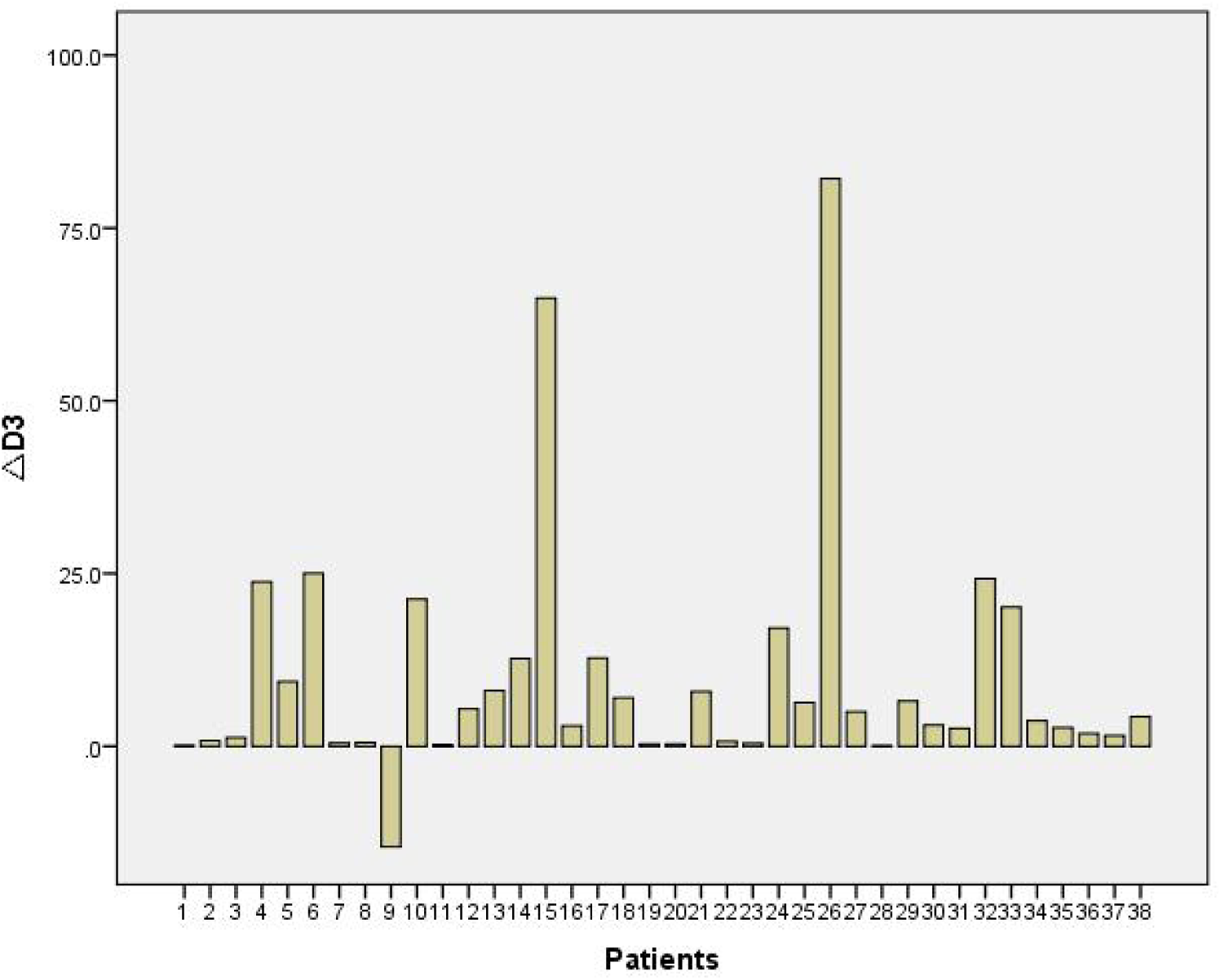
△D3: Difference in the D-dimer values between day 3 and day 1 after admission

**Figure 4.**
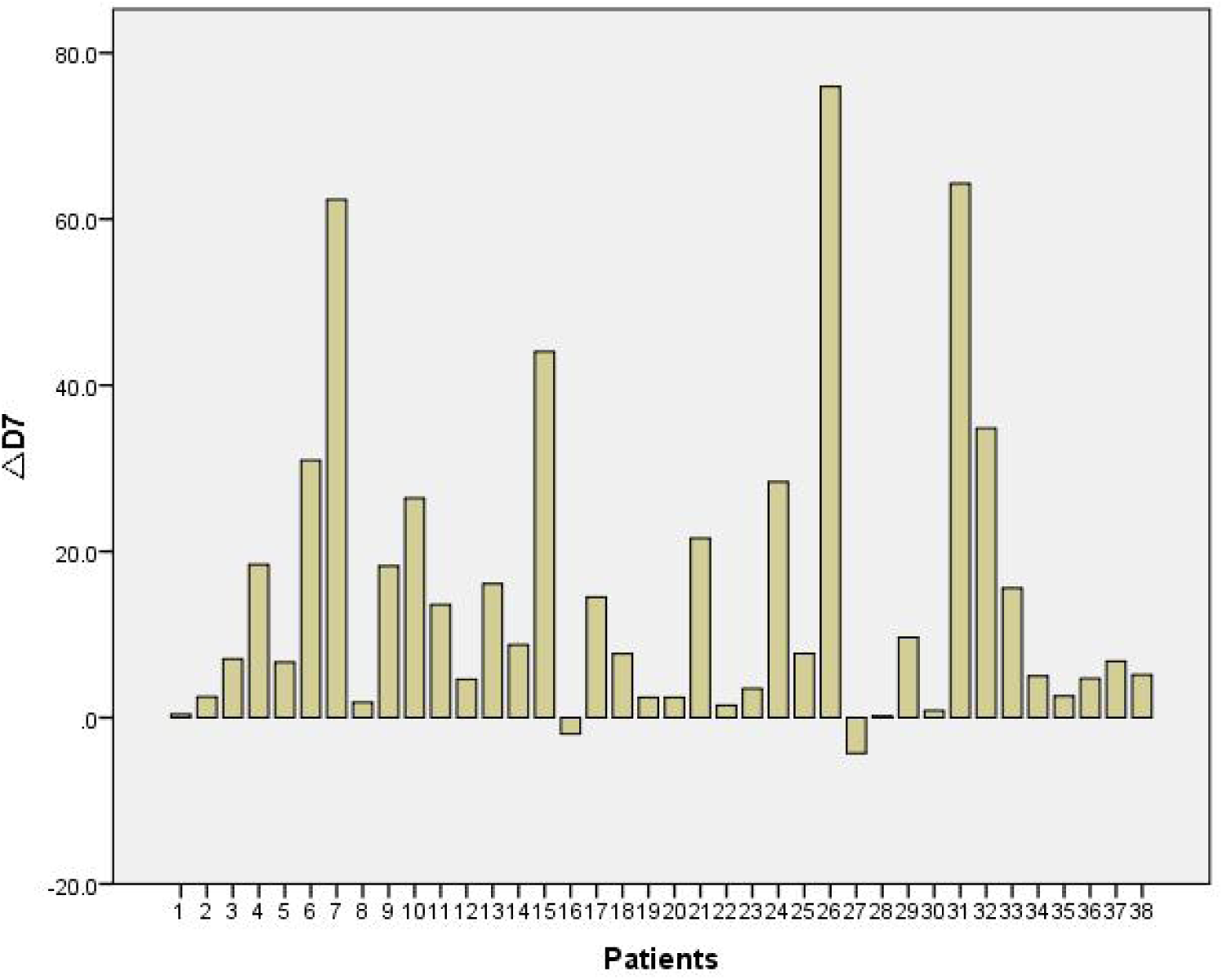
△D7:Difference in the serum D-dimer values between day 7 and day 1 after admission

**Figure 5:**
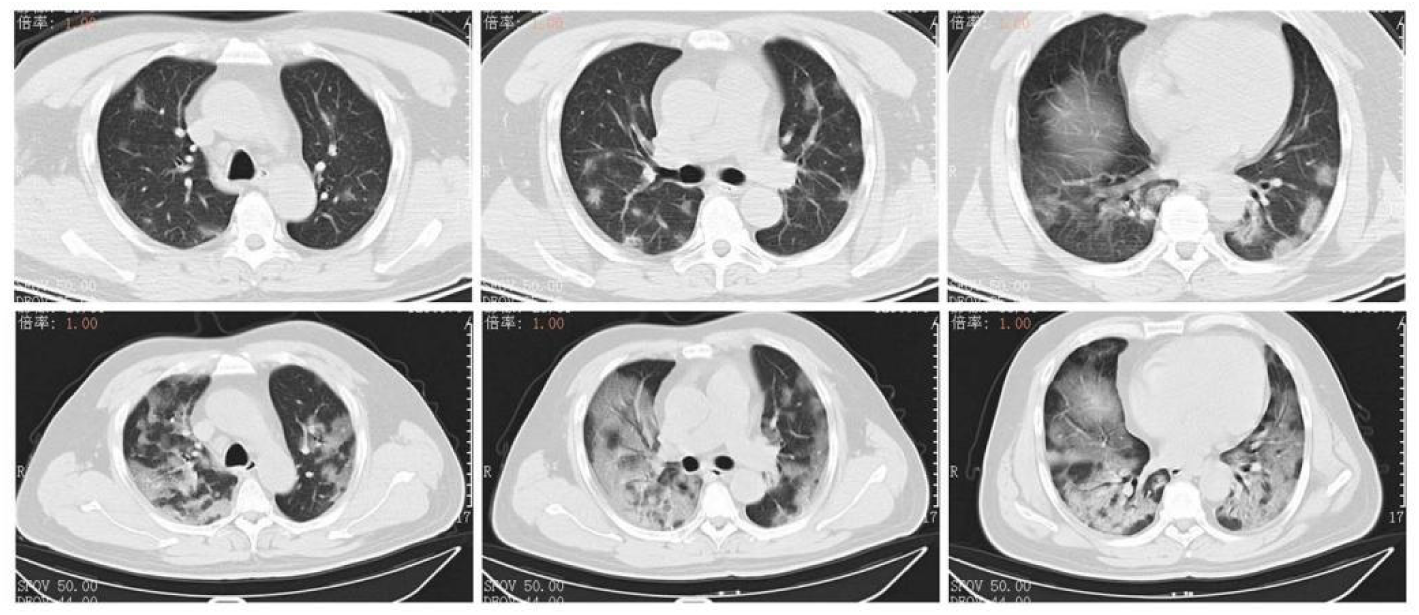
Top Row: chest CT obtained on Jan 10 (3A) showed mass shadows of the patchy glass in both lungs, which were distributed along the bronchial bundle and subpleurum. Next Row: Chest CT on Jan 13 showed improved status (3B) with diffuse consolidation of both lungs, uneven density and air bronchogram.

## Discussion

This was a cross-sectional study on the clinical characteristics of patients with 2019-nCoV, especially those with severe pneumonia. Our study suggested that advanced age, lymphocyte decline, and D-dimer elevation were more prominent in the patients with severe pneumonia, which is useful for the early identification of patients with severe pneumonia.

The laboratory examinations showed that patients with severe pneumonia had depressed serum albumin, elevated serum creatinine, blood urea nitrogen, aspartate aminotransferase, C-reactive protein, and B-type natriuretic peptide. Hypoproteinemia may be due to the patient’s consumption and inadequate protein intake caused by poor appetite. A previous study reported that hypoalbuminemia is a potent, dose-dependent predictor of poor outcomes for pneumonia with the coronavirus infection[11]. An elevated amount of C-reactive protein may be associated with the inflammatory response and cytokine storms caused by the virus in the blood vessels. These results were consistent with a previous study, which showed that the C-reactive protein level was positively correlated with the severity of the pneumonia[12].

According to the results of the binomial logistics regression analysis, we found that age and the level of the D-dimer were independent risk factors. These results suggested that the level of D-dimer was significantly positively correlated with the 2019-nCoV severe pneumonia, which was also shown in another study[13]. Previous studies showed that SARS-CoV could bind to ACE2, down-regulating the expressions of ACE2, and resulting in an increased Angiotensin II level in mouse blood samples, signaling through Angiotensin II receptor 1, and induced acute lung injury[14-16]. ACE2 is a receptor protein of both SARS-CoV and 2019-nCoV, and it is abundantly present in the epithelia of the lung and small intestine[17]. It was reported that 2019-nCoV binds to the ACE2 in the same way as SARS-CoV[18], inducing damage to the pulmonary arteries and leading to the extensive embolization in the extensive alveolar terminal capillaries. These changes eventually lead to an increase in D-dimer. A significant lymphocyte decline in the progression of severe pneumonia was also observed, which was consistent with the results of Huang et al[19]. Decreased lymphocytes suggested that 2019-nCoV may primarily attack the body’s immune system, especially the T lymphocytes, which is similar to the action of SARS-CoV[20]. After 2019-nCoV impairs the immune system, it is difficult to prevent the virus replication by immediately forming the neutralizing antibody. Non-specific pulmonary secondary inflammation acts with the 2019-nCoV infection, inducing cytokine storms and producing a series of immune responses and causing disorders of the lymphocyte subsets. Therefore, the above may explain the decline of lymphocytes and the rise of D-dimer in the progression of severe pneumonia. In addition, the pulmonary CT findings of patients with severe pneumonia can progress rapidly, so it is important to review chest CT scans in a timely fashion to learn about the pulmonary lesions.

### Limitations

This study had several limitations. First, only 110 patients from a single hospital were included in this study; a larger scale study needs to be carried out to confirm our conclusions. Second, most of the patients were still hospitalized when the manuscript was submitted, so we could not verify the efficacy of the therapeutic and prognosis of the patients.

## Conclusions

In general, the results suggested that advanced age, decreased lymphocytes, and elevated levels of D-dimer were risk factors for severe pneumonia. Clinicians should pay close attention to these indicators and identify high-risk patients as early as possible. More studies are needed to explore the clinical characteristics and treatment options of critically ill patients.

## Data Availability

The clinical data of the patients used in the study came from the central hospital of Wuhan and were approved by relevant departments of the hospital.Although the data included in the study included basic information about the patients, laboratory tests and imaging results, and had no privacy implications, the data could not be released at this time, as required by the hospital.

## Statements

### Acknowledgement

We thank LetPub (www.letpub.com) for its linguistic assistance during the preparation of this manuscript.

### Statement of Ethics

The study was approved by Ethics Committee of Wuhan Central Hospital (Yuan lun han [2020] no.4). As this study was a retrospective study, only clinical data of patients were collected, and privacy data such as name, ID number and telephone number were not involved, so no informed consent was obtained. Moreover, the data were only used for scientific research, not for other purposes.

### Author Contributions

Y.F.W. and Y.Z. had full access to all of the data in the study and take responsibility for the integrity of the data and the accuracy of the data analysis. Study concept and design: Y.F.W., S.G. Acquisition, analysis, or interpretation of data: Y.F.W., Y.Z., Z.Y., D.P.X.. Drafting of the manuscript: Y.F.W., Y.Z., S.G. Critical revision of the manuscript for important intellectual content: all authors. Statistical analysis: Y.F.W, Y.Z. Administrative, technical, or material support: D.P.X., Z.Y. Study supervision: S.G.

